# Dairy Environments with Milk Exposure are Most Likely to Have Detection of Influenza A Virus

**DOI:** 10.1101/2025.09.03.25335023

**Authors:** C. Stenkamp-Strahm, B. McCluskey, B. Melody, B. Christensen, N. Urie, N. Amey, R. Lomkin, A.J Campbell, S.S Lakdawala, J. Lombard

## Abstract

Highly pathogenic avian influenza virus of the H5N1 subtype has been infecting U.S dairy cattle and spreading among dairy farms since March 2024. H5N1 surveillance systems for dairy farms are needed, but information on whether environmental sampling can inform these systems is lacking. To guide a surveillance framework, we determined the environmental locations on H5N1-affected dairies (n = 25) in four states (California, Colorado, Michigan, and Ohio) that harbored influenza A virus (IAV), and explored sample characteristics that may influence viral detection. A total of 623 samples from environments and sale barns were characterized for IAV and classified into six categories based on location. A total of 94 samples (15.1%) had IAV detected, the majority in the following categories: milking equipment/personal protective equipment, parlor surfaces, and wastewater/lagoons/manure. These results suggest that dairy environments most likely to harbor IAV are those with exposure to milk, although the viral load in environmental samples was typically lower than that of bulk tank milk tested on a subset of farms. Mixed effect modeling was used to explore the relationship between IAV detection, Ct value, and days into an outbreak that samples were collected or the category where samples were collected. Days into an outbreak that samples were collected was associated with IAV detection while category of collection was associated with the measured Ct value. These results may guide H5N1 surveillance efforts on dairies, and can be strengthened by studies that collect samples from farm environmental locations prior to, during, and after H5N1 outbreak periods.

## INTRODUCTION

In February 2024, a disease syndrome characterized by fever, lethargy, dehydration and an abrupt drop in milk production affected cattle in dairy herds in Texas and Kansas. It was eventually confirmed that this syndrome resulted from an influenza A virus (IAV) of the H5N1 subtype (Burrough et al., 2024). Highly pathogenic avian influenza (HPAI) viruses are historically carried by wild birds, and are H5 or H7 subtype specific (CDC, 2025). Genetic investigations have deemed that reassortments in viral segments led to the Eurasian lineage goose/Guangdong clade 2.3.4.4b genotype B3.13, with a propensity for bovine hosts (Nguyen et al., 2024). The first bovine infection likely resulted from a spillover event from a wild bird having shed this virus, with subsequent cow-to-cow transmission occurring among herd mates. Since its discovery in cows, H5N1 of both the B3.13 and D1.1 genotypes have continued to spread in dairy cattle, with cows on dairy premises in 17 states affected (USDA, 2025a; USDA, 2025b).

During this outbreak, viral RNA has been regularly found in the milk but also variably found in the blood, nasal secretions and urine of clinically ill and nonclinical cows on H5N1-affected dairies (Caserta et al., 2024; Lombard et al., 2025a). Biosecurity recommendations include diverting the milk from affected cattle to waste streams, and treating this milk prior to discard (USDA, 2024c). During an outbreak, milk and other cattle excretions may serve to contaminate farm environments with H5N1 and propagate the virus to naïve cows within a herd or spread the virus elsewhere via fomites. To date, testing for IAV RNA in aggregate milk samples (i.e. bulk tank milk) from dairy herds has been leveraged for the surveillance and monitoring of H5N1-affected farms (USDA, 2024b). These surveillance efforts led to the discovery that the D1.1 genotype, in addition to the B3.13 genotype, had been introduced and was circulating in dairy cattle (USDA, 2025b). Given a continued need to keep milk from sick cows out of the saleable bulk tank, it is possible that samples other than bulk tank milk from H5N1-affected dairies are more sensitive for virus detection. Recently, wastewater surveillance in combination with bulk tank milk testing of dairies in the state of Oregon showed that environmental detections of virus were not likely of dairy origin (Sutton et al., 2025). Other methods to surveil dairies for H5N1, to the authors’ knowledge, have yet to be investigated.

IAV persistence in the environment outside of animal hosts has been previously studied. IAV has been shown to persist on several non-porous surfaces (steel, tile, rubber, plastic) for up to three days, and on latex for up to six days (Tiwari et al., 2006). A separate study indicated that under the right environmental conditions, H5N1 viruses can persist on certain substrates, such as galvanized metal, glass, and topsoil, for longer than 13 days (Wood et al., 2010). Environmental sampling conducted during previous HPAI outbreaks on poultry farms have shown viral detection from air samples (Torremorell et al., 2016) and different types of surfaces, including walls/doors, fans, cages, feeding equipment and gloves/masks (Lopez et al., 2018). Cattle-derived H5N1 strains have been detected and successfully isolated from both rubber and plastic on a farm in Kansas (Singh et al., 2024) and were seen to persist experimentally on stainless steel and rubber, the two substrates that comprise commercial milking equipment (Le Sage et al., 2024).

Environmental sampling for IAVs has been used within the poultry industries for early detection of HPAI outbreaks, identification of novel viral strains, and measuring success of outbreak intervention strategies (Hood et al., 2020). Environmental sampling has been explored for surveillance and the detection of IAVs in swine (Stadler et al., 2024), with environmental sampling showing good detection sensitivity (Garrido-Mantila et al., 2019). The utility of using environmental samples for surveillance within the dairy sector, either in lieu of or in addition to bulk tank milk sampling, is unknown, but environmental sampling methods may be convenient and cost-effective. Further, our understanding of how the H5N1 virus is transmitted within the farm environment remains limited (USDA, 2024a; Lombard et al., 2025b). Establishing which dairy farm environments harbor virus during farm outbreaks is advantageous, as it may guide future surveillance efforts and inform studies that further our understanding of H5N1 transmission on the dairy. Accomplishing this requires a survey of environmental samples taken from affected dairy operations in multiple geographic areas, at different points in their outbreak periods.

Here we describe detection of IAV RNA in environmental samples taken from multiple dairies affected by H5N1 in four states: California (CA), Colorado (CO), Michigan (MI) and Ohio (OH). We then use this information to guide consideration of environmental sampling for dairy H5N1 surveillance, and review the potential role of contaminated environments in H5N1 transmission.

## MATERIALS AND METHODS

### Study Design

The environmental samples analyzed in this work were collected as part of several separate studies and outbreak investigations. Figure 1 displays a timeline of when environmental and livestock market samples were collected, and when farms in each state experienced clinical signs of H5N1 disease in their cattle. The dates that dairy operations first experienced clinical signs were determined via conversation with herd owners/managers, private veterinarians, and via epidemiologic surveys. Supplemental Table 1 describes the size, cattle breeds and parlor types of participating dairies in each state. Specific details regarding study design and sample collection in each state are described below.

**Figure 1:**
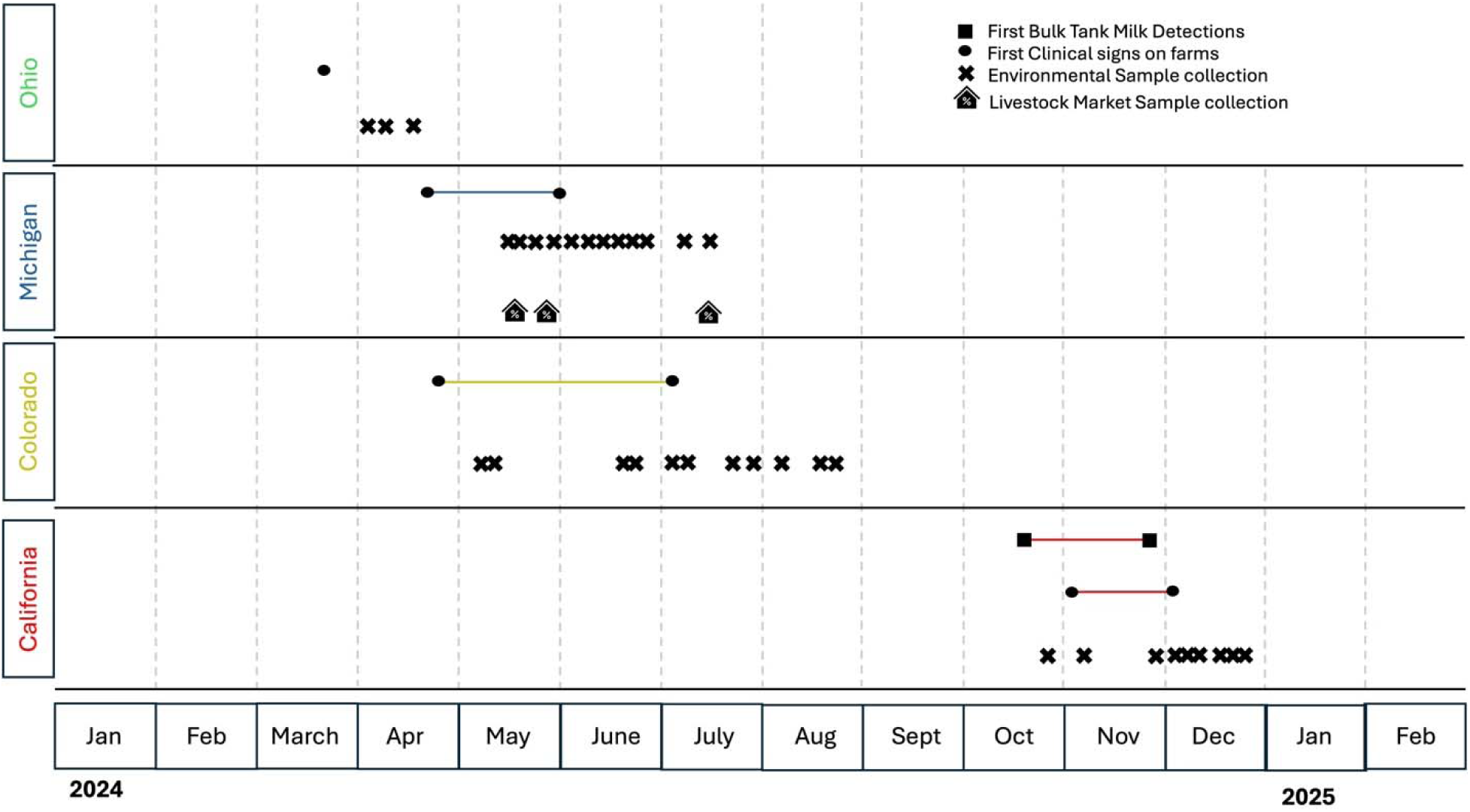
Timeline showing when study farms had initial clinical signs of H5N1 disease in cows, when bulk tank detections were first made (CA), and when samples were collected from livestock markets (MI) and dairy operations in each state.

In MI, H5N1 in cattle was first detected on March 29, 2024. A USDA Epidemiological Strike Team was invited by the Michigan Department of Agriculture and Rural Development (MDARD) to study affected dairy farms and identify potential links for viral dissemination. Twelve (12) operations and three sale barns in central and southern MI allowed collection of environmental samples from several locations on and surrounding their sites. These farms initially experienced clinical signs of the virus in spring 2024; between April 21 and May 31.

In OH, H5N1 in cattle was first detected on March 25, 2024. Samples were collected on a single dairy operation starting in early April 2024 by the Ohio Department of Agriculture (ODA) and the United States Department of Agriculture’s Animal and Plant Health Inspection Service, Veterinary Services (USDA:APHIS:VS). This operation experienced clinical signs of H5N1 in their cows on March 21, 2024.

In CO, H5N1 was first detected in cattle on April 21, 2024. The Colorado Department of Agriculture (CDA), USDA:APHIS:VS, and Colorado State University (CSU) recruited eight (8) affected premises in northeastern CO to participate in a study that aimed to collect multiple sample types from clinical and non-clinical cattle, over the course of multiple herd visits. On a subset of these premises (6) a variety of environmental samples were collected from sites on and surrounding each farm. These farms first experienced clinical signs of H5N1 in the late spring and summer 2024; between April 21 and July 7.

CDA mandated weekly bulk tank milk (BTM) testing of all commercial dairy herds within the state starting July 2024 (CDA, 2024). Ct values from weekly BTM samples from the herds that allowed environmental sampling (6) were provided to our team, with sample testing beginning July 29, 2024.

In CA, H5N1 was first detected in dairy cattle on August 30, 2024. In late September 2024 Lander Veterinary Clinic in Turlock, CA, recruited 19 non-affected dairy herds in central CA to begin voluntary daily BTM testing for H5N1, as part of a study designed to evaluate H5N1 detections in BTM over time. This effort was in collaboration with the California Department of Food and Agriculture (CDFA), USDA:APHIS, and CSU. All enrolled herds had virus detected between October 24 and November 28, 2024. On a subset of these farms (6), environmental samples were collected after BTM detection. Each farm experienced initial clinical signs of H5N1 in late fall to early winter 2024; between November 1 and December 1.

All premises enrolled in this study were confirmed as infected with IAV H5N1 clade 2.3.4.4b genotype B3.13 via testing at National Veterinary Services Laboratories (NVSL) in Ames, Iowa. These confirmations were made prior to environmental sampling for IAV.

### Environmental and BTM Sample Collection

Sterile polyester or cotton-tipped swabs, sterile gauze, or sterile swabs in pre-aliquoted universal viral transport media (UVT; Becton Dickinson, Cat. 220526) were used to sample various surfaces. Dry swabs and gauze were placed in 10-15ml transport media (brain heart infusion (BHI) or molecular transport media (MTM), provided by NVSL). Lagoon samples were collected by placing a sterile gauze pad or 50 mL conical tube on a weighted fishing rig, casting it into the lagoon, and allowing it to sink. Fluid collected from the saturated gauze or conical tube was then placed into another conical tube. All environmental samples were refrigerated if needed, and then either driven to the lab or shipped on ice overnight.

For the CA dairies, each operation submitted daily BTM samples starting at enrollment. Farms submitted between 1 and 8 of these samples per day, depending on operation size and daily milk production. An aliquot of each sample was tested for IAV at Lander Veterinary Clinic on the day of collection, and the remaining sample was shipped on ice overnight to the Iowa State University Veterinary Diagnostic Laboratory in Ames, IA.

The following National Animal Health Laboratory Network (NAHLN) laboratories completed BTM and/or environmental sample processing and testing: Michigan State University Veterinary Diagnostic Laboratory in Lansing, MI, Colorado State University Veterinary Diagnostic Laboratory in Fort Collins, CO, Iowa State University Veterinary Diagnostic Laboratory in Ames, IA, Washington Animal Disease Diagnostic Laboratory in Pullman, WA, Ohio Animal Disease Diagnostic Laboratory in Reynoldsburg, OH, and NVSL (Ames, IA). A subset of environmental samples from California dairies were transported and tested at Lander Veterinary Clinic in Turlock, CA.

### Sample processing, RNA extraction, rRT-PCR

Sample RNA was isolated and real-time reverse transcription polymerase chain reaction (rRT-PCR) assays for IAV were performed on environmental and BTM samples per NAHLN standard operating procedures and protocols, using a cycle threshold (Ct) cut-off value of 40. Annually, each NAHLN laboratory is required to successfully complete NVSL-administered proficiency tests and quality control procedures for identification of IAV. Virus isolation and whole genome sequencing (WGS) were not completed for any of the environmental samples collected in this study.

Environmental samples and initial BTM samples sent to Lander Veterinary Clinic were processed using the following protocol: UVT tubes were vortexed for 10 seconds with swabs inside and then centrifuged for roughly 10 seconds to collect liquid at the bottom of each tube. An aliquot was removed for RNA extraction and the remaining sample was stored at 4°C. RNA extraction was completed using 200-400ul of sample with a MagMax CORE Nucleic Acid Purification Kit (Applied Biosystems, Cat. A32700) paired with the Swine Influenza Virus RNA Test Kit (Applied Biosystems, Cat. 4415200) on a KingFisher Flex 96 system (ThermoFisher). A QuantStudio 5 Real-Time PCR System (ThermoFisher, Cat. A28569) was used according to manufacturer’s instructions to perform rRT-PCR for IAV. Reactions were performed using 8ul of extracted RNA from each sample.

### Analysis

BTM Ct values from several study farms in CA and CO were used to estimate the viral burden over time on operations experiencing H5N1 outbreaks. For CA farms, BTM Ct values were averaged by day. Ct values for each CO and CA farms were assigned based on the number of days between the BTM collection date and the date of first clinical signs in cows. BTM Ct values across all farms were averaged over time using a rolling 3-day mean.

Six environmental sample type categories were created to represent different sampling locations on affected farms. Personal protective equipment (PPE) was included in a category representing milking equipment, given its use during the milking process on each farm. Each sample type category and the number of samples collected are described in Table 1. Prior to exploring Ct results, characteristics and the distribution of samples collected was described. The number of samples in each category collected in each state was reported, and the number of days from clinical signs to sample collection was compared by state and sample category using ANOVA with Tukey’s post-hoc test (*p* < 0.05 as significant for comparisons).

**Table 1:** Summary of environmental samples for Influenza A virus detection by category and source, including number of samples collected.

| Sample Category | Sample source | Total samples collected |
| --- | --- | --- |
| Milking Equipment/PPE | Milking machine inflations | 100 |
|  | Water hoses | 25 |
|  | Gloves or apron of personnel | 20 |
|  | Milking machines | 9 |
|  | Dip equipment | 8 |
|  | Milk lines/hoses | 5 |
|  | Vacuum lines | 4 |
|  | Bulk tank filter/hose | 3 |
|  | Sinks | 3 |
|  | Total | 177 |
| Parlor Surfaces | Surfaces | 52 |
|  | Floor | 38 |
|  | Drain | 30 |
|  | Driveway next to parlor | 1 |
|  | Total | 121 |
| Wastewater/lagoon/manure | Lagoon/Manure | 39 |
|  | Lagoon Inflow | 12 |
|  | Wastewater/Flush water | 8 |
|  | Lagoon Outflow | 5 |
|  | Manure Spreader | 3 |
|  | Alley to Lagoon | 2 |
|  | Holding tank / pit | 2 |
|  | Total | 71 |
| Housing | Stanchion / Head lock | 49 |
|  | Alleyway / Driveway | 26 |
|  | Gates / Fences / Rails | 12 |
|  | Free Stall / Other Surface | 11 |
|  | Bedding | 7 |
|  | Fan / Shade Support | 3 |
|  | Barn Gutter | 1 |
|  | Total | 109 |
| Water tanks/feeders/feed | Waterers / tanks | 45 |
|  | Feeding equipment | 7 |
|  | Mineral block | 5 |
|  | Feed Bunk / Feeders / Feed | 5 |
|  | Total | 62 |
| Other | Sale barn equipment / surfaces | 42 |
|  | Farm equipment | 21 |
|  | Surfaces in rooms for personnel | 20 |
|  | Total | 83 |
| **Total samples** |  | **623** |

Ct values represent an estimate of the amount of IAV (viral load) present in each sample, and were reported by each NAHLN lab if values were less than the determined Ct cut-off of 40. Ct values from samples with and without IAV detection were described by sample category, state of sample collection, and the number of days from clinical signs to sample collection. For environmental sample categories with >1 IAV detection, Ct values from samples with detection were compared by category and state using ANOVA with Tukey’s post-hoc test (*p* < 0.05 as significant for comparisons).

Because several variables may influence IAV detection and the viral load (Ct value) of environmental samples, data were further explored via mixed regression modeling. The first aim was to understand the influence that the number of days after clinical signs a sample was collected, and the location a sample was collected from, had on IAV detection. To accomplish this, results from sample categories with >1 IAV detection were used in a mixed effect logistic regression model (Bates et al., 2015) using state of collection and laboratory as random effects, and days from clinical signs to sample collection or sample category as fixed effects. A second aim was to understand how the number of days after clinical signs that a sample was collected, and the location a sample was collected from, had on the viral load (Ct value) of samples with detection. Non-negative Ct values from sample categories with >1 IAV detection were tested for normality using a Shapiro-Wilk test. After log-transformation, mixed effect linear regression analyses were performed using state of collection and laboratory as random effects, and days from clinical signs to sample collection or sample category as fixed effects. P-values were determined for mixed models using likelihood ratio tests comparing full to null models. All statistical analyses were completed using R version 2024.12.1+563 or later.

## RESULTS

### Environmental Sampling Efforts on 25 Different Farms in Four States

After collecting environmental samples from 25 farms and three sale barns in four states, a total of 623 samples were characterized for IAV presence. We investigated the distribution of samples, given that these data were collected in separate studies that had different aims. The number of samples collected from each category varied by state (Supplementary Figure 1). The days from clinical signs that samples were collected varied by sample category, with wastewater/lagoon samples being collected later in a farm outbreak period compared to all other categories (*p* < 0.001 for all comparisons), and milking equipment/PPE samples being collected a greater number of days since initial clinical signs compared to samples taken from parlor surfaces (*p* < 0.001) and water tanks/feeders/feed (*p* = 0.004) (Supplementary Figure 2A). The days from clinical signs that samples were collected also varied by state, with CA samples, on average, being collected earlier in a farm outbreak period compared to other states, and MI samples, on average, being collected later in a farm outbreak period compared to other states (*p* < 0.001 for all comparisons; Supplementary Figure 2B). For Michigan, sample collection ranged from 6 to 64 days after first clinical signs with a mean of 35 days. The Ohio farm had samples collected from 15 to 29 days after first clinical signs with a mean of 22 days. Colorado operations had samples collected 10 to 69 days after first clinical signs with a mean of 21 days. Lastly, California farms had samples collected 3 days prior to first clinical signs to 21 days after first clinical signs with a mean sampling time of 3 days after clinical signs.

### Detections of Influenza A were Made in Environments Likely to be Contaminated with Milk

The main aim of this investigation was to understand where IAV could be found in the environment of H5N1-affected dairies. A total of 94 (15.1%) of the 623 environmental samples collected had IAV detected (Table 2). The largest percentage of positive samples were detected on parlor surfaces (28.9%) which had the second largest number of samples collected for a single category (n=121) and represented 19.4% of all samples collected. There were 26.6% of milking equipment and PPE samples that were positive, and they represented 28.4% of all samples collected. Meanwhile, 14.1% of wastewater/lagoon/manure samples were positive. Only a single sample from the housing category (an alleyway sample from MI), and one sample from the other category (a sample taken from equipment used to give cows liquid treatments in OH), were positive. This represents about 1% of samples tested in each of those categories. No samples collected from sale barns (n=42), or the water tanks/feeders/feed category (n=62) had IAV detected. Ohio had the largest percentage of samples test positive at 27.1% (13/48) followed by CA at 21.6% (43/199). Colorado had 11.1% samples test positive (26/235) followed by MI at 8.5% (12/141).

**Table 2.** Summary of environmental sample Influenza A virus PCR-positive test results, by sample category and state of collection.

| Sample category | PCR Positive Samples by State |  |  |  |  |  |  |  |  |  |
| --- | --- | --- | --- | --- | --- | --- | --- | --- | --- | --- |
|  | CA |  | CO |  | MI |  | OH |  | Category Total |  |
|  | Number | Percent | Number | Percent | Number | Percent | Number | Percent | Number | Percent |
| Milking Equipment/PPE | 19/46 | 41.3% | 21/106 | 19.8% | 0/1 | 0.0% | 7/17 | 41.2% | 47/177 | 26.6% |
| Parlor Surfaces | 23/60 | 38.3% | 5/39 | 12.8% | 2/6 | 33.3% | 5/16 | 31.3% | 35/121 | 28.9% |
| Wastewater/lagoon/manure | 1/4 | 25.0% | 0/2 | 0.0% | 9/65 | 13.8% | 0/0 | 0.0% | 10/71 | 14.1% |
| Housing | 0/44 | 0.0% | 0/37 | 0.0% | 1/26 | 3.8% | 0/2 | 0.0% | 1/109 | 0.9% |
| Water tanks/feeders/feed | 0/29 | 0.0% | 0/30 | 0.0% | 0/1 | 0.0% | 0/2 | 0.0% | 0/62 | 0.0% |
| Other | 0/16 | 0.0% | 0/21 | 0.0% | 0/42 | 0.0% | 1/4 | 25.0% | 1/83 | 1.2% |
| **Total** | 43/199 | 21.6% | 26/235 | 11.1% | 12/141 | 8.5% | 13/48 | 27.1% | 94/623 | 15.1% |

### Viral load of IAV in Environmental Samples was Typically Lower than that of BTM

Infected cows shedding virus likely contribute to the detection of virus in farm environments. Because of this, we performed a visual comparison of detections in environmental samples and BTM samples over the course of a farm outbreak period. Samples with no detection were given a value of 40 and the distribution of Ct values was visualized by category, state, and days after clinical signs that each sample was collected (Figure 2). In general, lower Ct values were seen from samples collected earlier in farm outbreak periods, and Ct values from samples taken over time paralleled or were higher than average BTM Ct values used to designate the viral burden on a farm (Figure 2).

**Figure 2:**
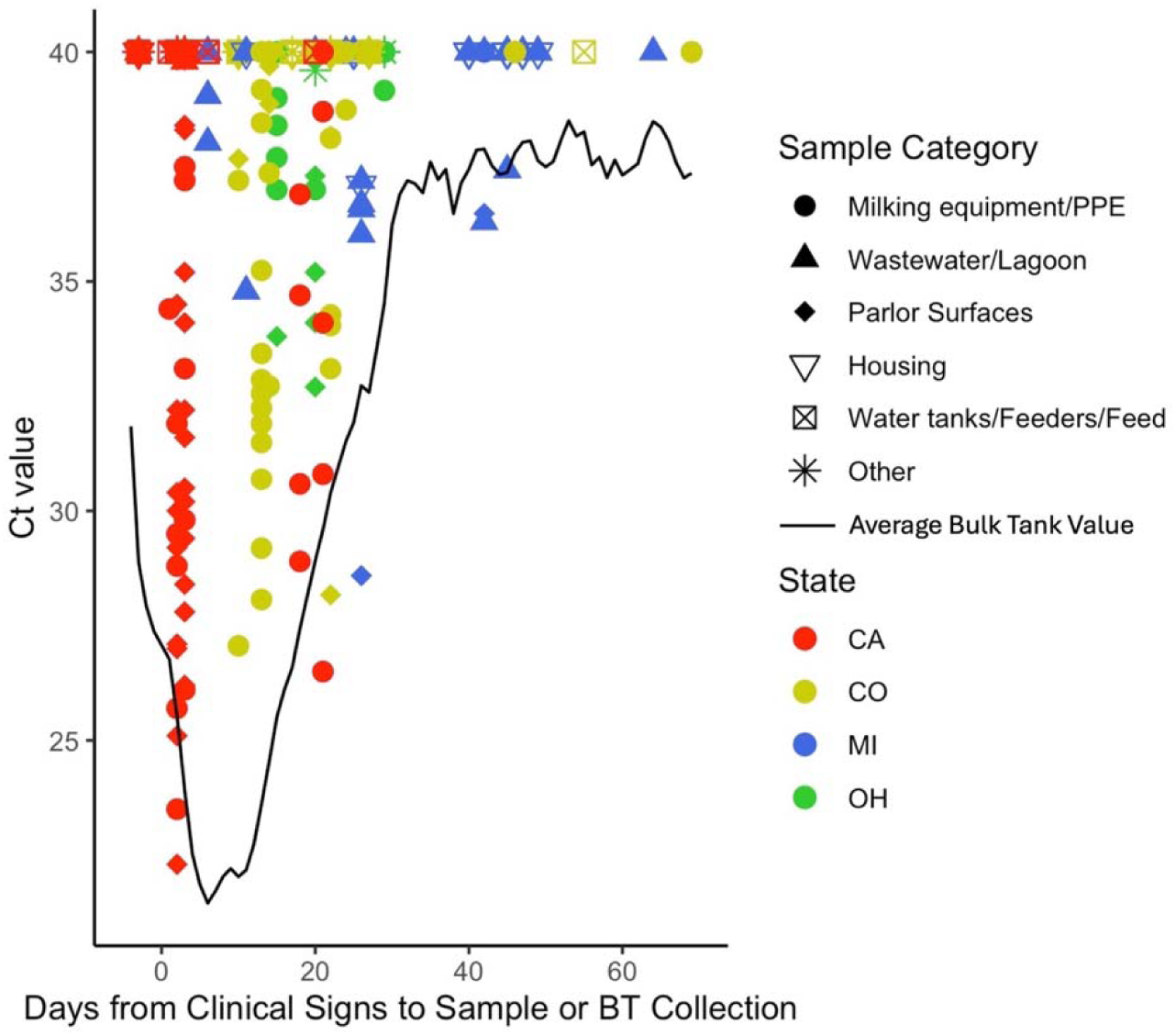
Ct values of environmental samples by state of collection and days from clinical signs to collection, overlaid with a curve of averaged bulk tank (BT) milk Ct values acquired from 10 study farms during the period of sampling. Ct values < 40 were considered to have IAV detected, samples classified as negative have been given a value of 40.

### Of Locations with IAV Detection, Highest Viral Loads were Seen from Milking Equipment, Parlor Surfaces

We wanted to understand what environmental samples with IAV detection had the highest viral loads (lowest Ct values), as these locations may influence viral transmission or be more successfully utilized in future H5N1 surveillance efforts. The average Ct values for samples with IAV detection were greater than 30 (Table 3), with the lowest Ct values in samples from parlor surfaces (avg. 31.9 +/− 4.5 cycles) and milking equipment/PPE (avg. 33.5 +/− 4.3 cycles). The average Ct value of PCR-positive wastewater/lagoon samples was significantly higher than parlor surface sample categories (*p* = 0.002) and milking equipment/PPE (*p* = 0.035; Figure 3A), and the average detected Ct values from positive samples were also significantly different by state, with CA having lower average Ct values compared to CO (*p* = 0.006), MI (*p* = 0.001) and OH (*p* < 0.001; Figure 3B).

**Figure 3:**
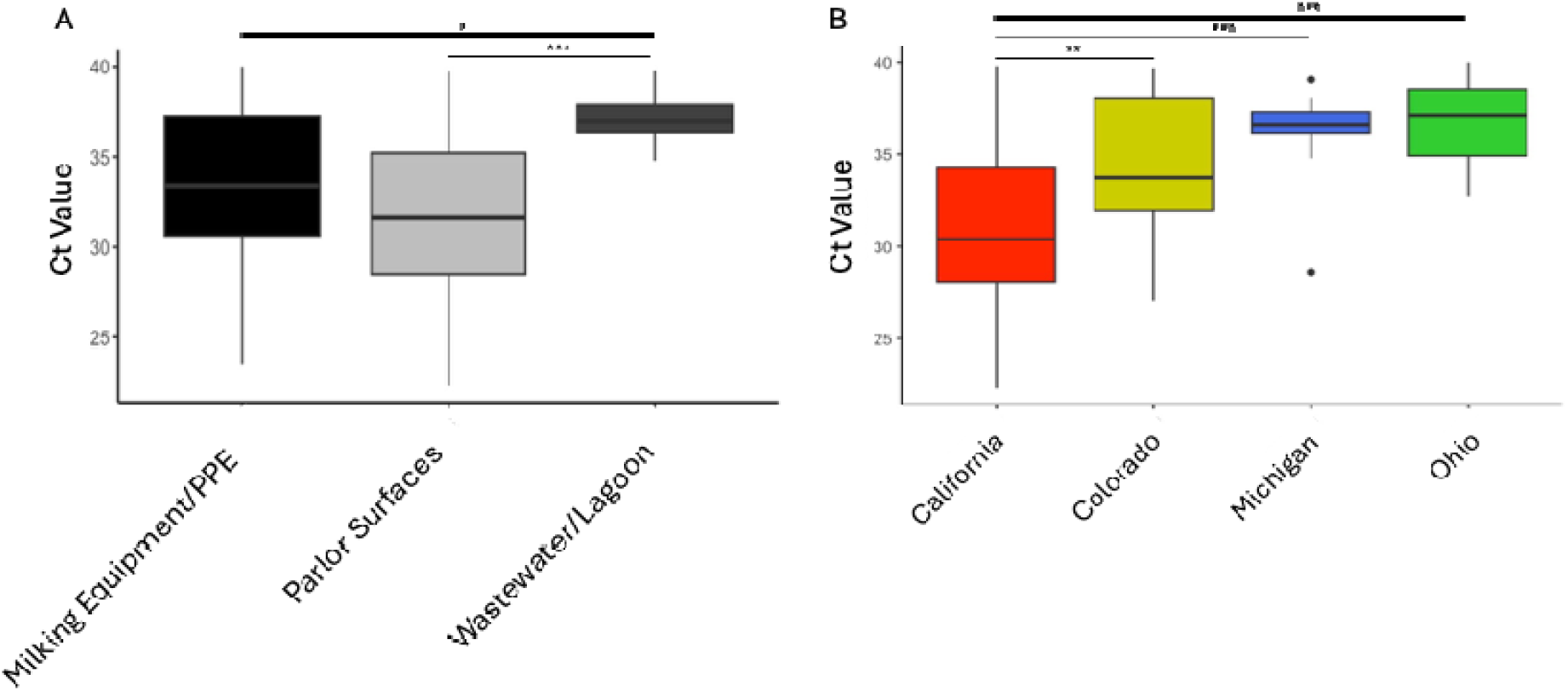
Panel A: The range of Ct values of environmental samples collected for Influenza A virus with detection (Ct<40) via PCR testing by sample source category. Panel B: The range of Ct values of environmental samples collected for Influenza A virus with detection (Ct<40) via PCR testing by state of collection. For statistical significance: * *p* < 0.05, ** *p* <0.01, *** *p* <0.001.

**Table 3:** The average and range of Ct values measured in the five environmental sample source categories with Influenza A virus detected (Ct<40) via PCR testing.

| Sample source category | Average positive sample Ct value (SD) | Range of Ct values |
| --- | --- | --- |
| Milking Equipment/PPE | 33.5 (4.3) | 23.5 – 39.9 |
| Parlor Surfaces | 31.9 (4.5) | 22.3 – 39.7 |
| Wastewater / lagoon / manure | 37.2 (1.5) | 34.8 – 39.8 |
| Housing | 37.1 | 37.1 |
| Other | 39.6 | 39.6 |

A total of 55 samples (8.8%) had indeterminate or quality flagged Ct results (data not shown) and were classified as negative. Environmental samples often contain matter that inhibits the rRT-PCR assay, which causes indeterminate results (Schrader et al., 2012). Indeterminate or quality flagged samples were collected in CO and MI, and were from several sample type categories.

### Odds of Detecting IAV in Environmental Samples Decreased Over Outbreak Course

Because the goal of the current analysis was to generalize where IAV may be found in the dairy farm environment, and samples had been collected in a non-uniform manner across states, epidemiologic modeling was used to understand the influence of certain variables on results. In addition to the sampling location, several factors may play a role in reported IAV detections including the number of days that sampling occurred relative to the start of the outbreak in the herd (first clinical signs), the states samples were collected in, and the laboratories that tested each sample. In this analysis, the state of collection also represents a season, given when each state experienced H5N1-associated disease. The laboratories doing the testing of samples were associated with the state of sample collection; two different NAHLN laboratories tested samples from each state, and there was very little overlap between laboratories and states. The farms enrolled were also correlated with laboratory and state, as each farm (with two exceptions) submitted samples to only one laboratory. We used mixed effect modeling to control for the effect of state and laboratory, as these were not uniform across sample type categories and days to collection, and initial Ct values had showed variability by state.

For the mixed effect logistic regression modeling, individual sample results from categories with >1 IAV detection were categorized as 0 / 1 (non-detected / detected) and used as the dependent variable. Laboratory was nested within state as a random effect, and days from clinical signs to sample collection and sample category were modeled as fixed effects. When compared to the null model, category of sample collection, when used as a fixed effect alone, was not associated with IAV detection (*p* = 0.246) (Table 4). The number of days from first clinical signs that samples were collected, as a fixed effect alone, was associated IAV detection (*p* = 0.0015; OR 0.96 (0.91-0.98), indicating that as the number of outbreak days a sample is collected increases, the odds of detecting IAV in that sample decreases (Table 4). When a mixed logistic model was created using both fixed variables of interest (sample category, days from clinical signs that samples were collected), days from clinical signs was still associated with IAV detection and its effect estimate was relatively unchanged (data not shown).

**Table 4:** Results of mixed effect logistic regression models evaluating the association between IAV detection and sample category or days from clinical signs that samples were collected.

| Variable | Level | Fixed Effects |  |  | Random Effects |  |
| --- | --- | --- | --- | --- | --- | --- |
|  |  | Estimate (SE) | P-value<sup>1</sup> | Odds Ratio (CI) | Lab SD | State SD |
| Intercept |  | -1.62 (0.70) | 0.246 |  | 1.08 | 0.00 |
| Sample Category | Wastewater/Lagoon | Referent |  |  |  |  |
|  | Milking Equipment/PPE | 0.85 (0.67) |  | 2.32 (0.62 – 9.10) |  |  |
|  | Parlor Surfaces | 1.06 (0.65) |  | 2.88 (0.80 – 10.67) |  |  |
| Intercept |  | 0.148 (0.51) | 0.0015 |  | 0.96 | 0.00 |
| Days from Clinical Signs to Sample Collection |  | -0.052 (0.018) |  | 0.95 (0.91 – 0.98) |  |  |

<sup>1</sup> Maximum likelihood estimation comparing specified model to a null model using only random effects

### The Location of Sample Collection Influenced Viral Load

For the mixed effect linear regression, positive results from categories with > 1 IAV detection were evaluated using Shapiro-Wilk (*p* = <0.001) and the Ct values were determined to have a non-normal distribution. Ct values were then log-transformed and the model was constructed using a random effect of laboratory nested within state. When compared to the null model, category of sample collection, when used as a fixed effect alone, was associated with detected Ct value (*p* = 0.037) (Table 5), while the number of days from first clinical signs that a sample was collected, as a fixed effect alone, was not (*p* = 0.20) (Table 5). When a mixed model was created using both fixed variables of interest (sample category, days from first clinical signs that samples were collected), category was still predictive of detected Ct value and its effect estimate was unchanged (data not shown). These linear model outcomes suggest that the location of sample collection is more influential on the measured Ct value than the number of days into an outbreak that the sample was collected.

**Table 5:** Results of mixed effect linear regression models evaluating the association between the log-transformed Ct value of positive samples and sample category or days from clinical signs that samples were collected.

| *Variable* | *Level* | *Fixed Effects* |  | *Random Effects* |  |
| --- | --- | --- | --- | --- | --- |
|  |  | Estimate (SE) | P-value<sup>1</sup> | Lab SD | State SD |
| Intercept |  | 3.67 (0.032) | 0.037 | 0.049 | 0.027 |
| Sample Category | Wastewater/Lagoon | Referent |  |  |  |
|  | Milking Equipment/PPE | -0.053 (0.027) |  |  |  |
|  | Parlor Surfaces | -0.065 (0.026) |  |  |  |
| Intercept |  | 3.62 (0.03) | 0.20 | 0.046 | 0.027 |
| Days from Clinical Signs to Sample Collection |  | 0.0006 (0.0005) |  |  |  |

<sup>1</sup> Maximum likelihood estimation comparing specified model to a null model using only random effects

## DISCUSSION

The current study describes environmental locations on 25 H5N1-affected dairy farms that had IAV viral RNA detection. These survey results not only describe IAV detections on a large number of farms for the first time, but they also provide a robust framework for future studies assessing dairy environmental sampling as an IAV surveillance tool.

Most IAV detections were made in samples collected from three location categories: milking equipment/PPE, parlor surfaces, and wastewater/lagoons. If individual cow or BTM sampling were not feasible on a farm and confirmation of H5N1 infection were needed, the information in this work can be used to target sampling of these specific locations where IAV would likely be detected. To the authors’ knowledge, only one other study has assessed contamination of the environment on a dairy farm experiencing H5N1, and although this study showed detections from different substrates (rubber, plastic), it did not provide information on specific locations or the number of days from first clinical signs that samples were collected (Singh et al., 2024). Our results include samples taken from many operations at different stages of outbreak, and suggest that IAV is more likely found in farm environments that have direct contact with milk. This hypothesis is supported by the relatively low Ct values from viral RNA (i.e., high viral load) detected in contaminated milk, although different cattle excretions containing viral RNA (e.g., urine, nasal discharge, blood) (Lombard et al., 2025a, Caserta et al., 2024) may also contaminate these locations. It is also possible that the ‘micro’ environments with IAV detection (e.g., milking equipment, parlor surfaces, lagoons) are more conducive to IAV viral persistence than the other locations tested. Studies have shown that IAV may persist better in environments with variable humidity, which are cool and away from sun exposure (Brown et al., 2009; Sagripanti and Lytle, 2007; Lowen et al., 2007), although this may depend on viral subtype and attributes of the host shedding the virus (Kormuth et al., 2019; Kormuth et al., 2018; Qian et al., 2023). Other factors that influence viral persistence include droplet size and evaporation rates, open versus closed air environments, and the porosity of surfaces IAV is deposited on (French et al., 2023; Weber and Stilianakis, 2008). The current analysis also highlights the presence of IAV on PPE samples taken from workers; 4/47 positive samples from the milking equipment/PPE category were those collected from worker gloves and a worker apron.

Our data show lower Ct values in milking equipment and parlor surface samples compared to lagoon samples, supported by mixed effect linear regression modeling that controlled for state and testing lab. One may expect viral loads from wastewater streams to be dilute compared to those of surfaces in the parlor, supporting this finding. Although PCR detection does not differentiate between live virus and non-viable viral RNA, virus isolation and whole genome sequencing (WGS) is often more successful in samples with higher viral loads (Latorre-Margalef et al., 2016; Horm et al., 2016). The current study did not attempt to isolate live virus or perform WGS on collected samples, given both the time between sample collection and laboratory testing, and the magnitude of Ct values from many of the samples that had IAV detected. If future environmental sampling is done with a goal to perform virus isolation or WGS, targeting a location (e.g., parlor surface) that is more likely to have a Ct < 30 should be pursued. Historically, virus isolation rates from environmental samples collected during avian studies have been variable, likely due to the amount of viable virus in samples, sample processing and laboratory techniques (Haynes et al., 2009; Onuma et al., 2017; Vong et al., 2008; Grillo et al., 2015; Horm et al., 2016). Whether or not dairy environmental samples can support phylogenetic surveillance for H5N1 will hinge on future studies that assess both the viral load of collected samples, and the suitability of currently available viral culture techniques.

A relatively low percent (15.1%) of environmental samples had IAV detected in the current study. Sample collection methods and the distribution of samples available was influenced by the design of specific studies performed in each state. Mixed effect logistic regression modeling that controlled for state and testing lab showed that IAV detection was influenced by the time into an outbreak that samples were collected. This supports a notion that as an outbreak wanes and cattle recover from H5N1 disease, their viral shedding decreases resulting in a decrease of virus in the environment. BTM results from 10 sample dairies were used to represent the relative load of H5N1 in the farm environment during the same outbreak time-period that environmental samples were collected, and the Ct values from both sample types increased in a similar fashion. Viral loads from environmental samples taken at similar time points were also near or lower than those of BTM, backing the concept that excretions of cows shedding virus have a large impact on environmental IAV detection. These results and those of mixed modeling suggest that environmental sampling for IAV detection may be most successful during the farm outbreak peak. However, BTM information was not available for all study farms, and 1/10 dairies that supplied data had continued BTM Ct detection of IAV (Ct < 40) through the end of the 70-day study window. Relatively few samples were collected beyond 40 days from clinical signs in the current work, and the single farm with continued BTM detection was sampled one time very early in their outbreak period. Results may have been different were this farm sampled later, or if persistent sampling of other farms in the current study had been performed. Future work to assess the utility of environmental sample collection should consider collecting samples at multiple time points on the same farm, and perform sampling on farms with continued or chronic BTM detections.

Environmental sampling of live bird markets has been used for early detection of H5N1 (Cauthen et al., 2000). Because dairy environmental samples may be collected with ease compared to those from individual cattle, there is use in knowing if these samples could be used for early detection of H5N1-affected farms. One CA operation in the current study had environmental samples collected from multiple location categories prior to cattle showing clinical signs of disease. All environmental samples collected on this farm in this period did not have IAV detected, and BTM samples collected during the same window had Ct values as low as 23.7. Although sampling this early was only performed on one farm, these results suggest that aggregate or BTM sampling may be superior for early detection of H5N1 affected dairy farms. This conclusion would be strengthened by additional environmental sampling on farms prior to clinical signs in cattle.

Importantly, it is unknown if the collection of other types of samples (e.g., bioaerosol samples or transient fomites like flies, peridomestic species, humans) from the early sampled farm and the other farms in this study would have yielded the same results. Detection of IAV in air samples from porcine operations have been highly correlated with detection in environmental and individual animal samples (Prost et al., 2018). During the current dairy outbreak, IAV was unable to be detected in air samples collected on two H5N1 affected dairy operations in Texas (Shittu et al., 2024). However, detections were made in aerosols collected from affected dairy operations in California (Campbell et al., unpublished data) and in a Vaccine and Infectious Disease Organization Containment Level 3 (VIDO CL3) large animal facility where cows were experimentally infected via the intramammary route (Facciuolo et al., 2024). Because shedding of the H5N1 virus into the air by experimentally infected ferrets has been correlated with transmissibility (Tosheva et al., 2024), future environmental work should also include the collection of bioaerosols from naturally infected herds. Prospective sampling of bioaerosols from naïve dairy herds in affected states may also be beneficial.

### Limitations

The current study had several limitations. The time between sample collection and laboratory testing did not allow for targeted sampling of environmental locations that had IAV detection. And although our data set was large and represented multiple farms, the number of samples from each category collected from each farm differed, and total samples from each category varied over the outbreak period. Although it is one of the only markers available, using the time from first clinical signs as a temporal indicator of when a farm outbreak starts has inherent limitations also, as clinical signs and detection thereof can vary among farms. As mentioned, humidity, temperature, exposure to solar radiation, and other factors contribute to the persistence and viability of IAVs in the environment (Keeler et al., 2014; Brown et al., 2007; Sagripanti and Lytle, 2007; Lowen et al., 2007). MI and OH, CO, and CA experienced H5N1 infection in dairy cattle during spring, summer, and fall, respectively, so sampling of farms in each of these states reflected weather patterns and seasonal variation in some of these parameters. The goal of the current analysis was to generalize where IAV may be found in the dairy farm environment, regardless of season and geographic location, to better inform environmental sampling as a dairy surveillance tool. We therefore aimed to control for the effect of state and laboratory via mixed effect modeling. The distribution of collected samples may have muted the effect that days from first clinical signs to sample collection had on detected Ct values in our linear model, given that variation by state was controlled for. Future work assessing dairy environmental locations that harbor IAV should aim to consistently collect samples from farm locations prior to, during, and after their cattle experience clinical signs of disease. This would allow a temporal understanding of how the virus persists in certain sites on the dairy and, if appropriately powered, allow an assessment of which farm management factors influence the persistence of virus at these sites. Since the H5N1 virus is a pathogen only recently impacting the dairy industry, we also do not have a great understanding of how sampling technique influences the detection sensitivity for virus in these environments. Methods used to sample poultry environments have been tested and refined since the 1970s (Hood et al., 2020), with studies showing that different sample pre-processing, elution, and concentration methods improve IAV detection in different scenarios (Horm et al., 2011; Deboosere et al., 2011; Ronnqvist et al., 2012). The presence of inhibitors or other sample attributes may have affected IAV detection sensitivity, causing some of the indeterminate results seen in this study. Future work should explore how different sampling methods, especially the use of pre-wetted swabs (Julian et al., 2011), impact viral recovery and detection in samples taken from dairy surfaces.

## Conclusion

Since detection in dairy cattle in early 2024, dairy surveillance for IAV has become a salient topic. The success of environmental sampling for IAV surveillance in any industry is dependent on whether the ultimate goals are detection of virus, phylogenetic analyses of viral strains, monitoring viral burden over time, or monitoring viral burden after initiating an intervention. Samples and results collected for this study provide a strong framework within which to continue the assessment of environmental sampling as a surveillance tool to manage and mitigate dairy H5N1, especially as we learn more regarding viral shedding and transmission of the virus among dairy cattle.

## Data Availability

All data produced in the present study are available upon reasonable request to the authors

## Acknowledgements

The authors would like to acknowledge the following NAHLN labs for their work characterizing samples: Michigan State University Veterinary Diagnostic Laboratory, Colorado State University Veterinary Diagnostic Laboratory, Iowa State University Veterinary Diagnostic Laboratory, Washington Animal Disease Diagnostic Laboratory, Ohio Animal Disease Diagnostic Laboratory, and the National Veterinary Services Laboratory. We would like to additionally thank the state level partners (California Department of Food and Agriculture, Colorado Department of Agriculture, Michigan Department of Agriculture and Rural Development and Ohio Department of Agriculture) for their collaboration and cooperation during completion of this work. The authors would lastly like to thank the following personnel of Lander Veterinary Clinic for their work collecting and characterizing samples: Yvette Guzman, Giselle Leon, Ernesto Padilla, Alondra Ramirez, Carina Montanez, and Jessica Pacheco.

