## Supplemental Tables and Figures for "Dairy Environments with Milk Exposure are Most Likely to Have Detection of Influenza A Virus"

**Supplemental Table 1:** Demographic characteristics of 25 herds enrolled for environmental sampling, by State

|  | | MI | | OH | | CO | | CA | |
| --- | --- | --- | --- | --- | --- | --- | --- | --- | --- |
| **Characteristic** | **Level** | Number | Percent | Number | Percent | Number | Percent | Number | Percent |
|  | Total | 12 | 100.0 | 1 | 100.0 | 6 | 100.0 | 6 | 100.0 |
| Herd Size | <100 | 0 | 0.00 | 0 | 0.0 | 0 | 0.00 | 0 | 0.00 |
|  | 100 - 499 | 3 | 25.0 | 0 | 0.00 | 0 | 0.00 | 0 | 0.00 |
|  | 500+ | 9 | 75.0 | 1 | 100.0 | 6 | 100.0 | 6 | 100.0 |
| Parlor Type | Herringbone | 9 | 75.0 | 0 | 0.00 | 2 | 33.3 | 2 | 33.3 |
|  | Parallel | 1 | 8.33 | 1 | 100.0 | 4 | 66.7 | 1 | 16.7 |
|  | Rotary | 0 | 0.00 | 0 | 0.00 | 0 | 0.00 | 2 | 33.3 |
|  | Robotic | 1 | 8.33 | 0 | 0.00 | 0 | 0.00 | 1 | 16.7 |
|  | NA* | 1 | 8.33 | 0 | 0.00 | 0 | 0.00 | 0 | 0.00 |
| Breeds | Holstein >= 95% | 10 | 83.3 | 0 | 0.00 | 3 | 50.0 | 5 | 83.3 |
|  | Jersey >= 95% | 0 | 0.00 | 0 | 0.00 | 0 | 0.00 | 1 | 16.7 |
|  | Mixed | 2 | 16.7 | 1 | 100.0 | 3 | 50.0 | 0 | 0.00 |

*Refers to dairy with parlor at other site location

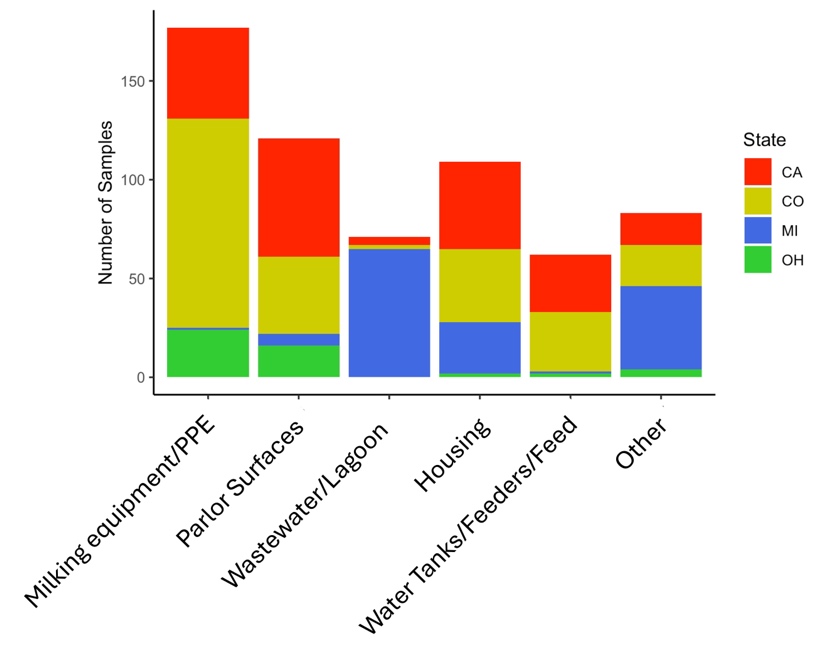

**Supplemental Figure 1:** Number of environmental samples collected by sample source category and by state for Influenza A virus detection.

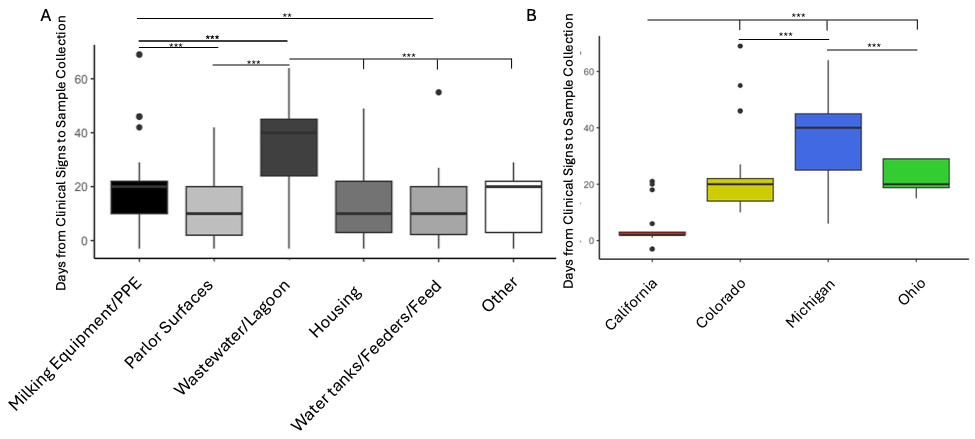

**Supplemental Figure 2:** Panel A: Boxplots of the days from clinical signs to environmental sample collection for Influenza A detection by sample source category. Panel B: Boxplots of the days from clinical signs of Influenza A to environmental sample collection by state of collection. For statistical significance: * *p* < 0.05, ** *p* <0.01, *** *p* <0.001.
